# Re-irradiation to the Prostate using stereotactic body radiotherapy (SBRT) after initial definitive Radiotherapy – A systematic review and Meta-analysis of recent trials

**DOI:** 10.1101/2024.03.05.24303777

**Authors:** Christina Schröder, Hongjian Tang, André Buchali, Daniel Rudolf Zwahlen, Robert Förster, Paul Windisch

## Abstract

**Background:** There is increasing data on re-irradiation to the prostate using stereotactic body radiotherapy (SBRT) after definite radiotherapy for prostate cancer. There has been increasing evidence on prostate re-irradiation using a C-arm LINAC or a MR LINAC in the last years. We therefore conducted this systematic review and meta-analysis on prostate re-irradiation including studies published from 2020-2023 to serve as an update on existing meta-analysis.

**Methods:** We searched the Pubmed and Embase databases in October 2023 with queries including combinations of “repeat”, “radiotherapy”, “prostate”, “re-irradiation”, “reirradiation”, “re treatment”, “SBRT”, “retreatment”. Publication date was set to be from 2020 to 2023. There was no limitation regarding language. We adhered to the Preferred Reporting Items for Systematic Reviews and Meta-Analyses (PRISMA) recommendations. After data extraction, heterogeneity testing using I2. Afterwards a random effects model with a restricted maximum likelihood estimator was used for estimating the combined effect. Funnel plot asymmetry was assessed visually and using Eggers test to estimate the presence of publication and/or small study bias.

**Results:** 14 publications were included in the systematic review. The rates of acute ≥ grade 2 (G2) GU and GI toxicities reported in the included studies range from 0.0-30.0% and 0.0-25.0% respectively. For late ≥G2 GU and GI toxicity, those values are 4.0-51.8% and 0.0-25.0%. The pooled rate of acute GU and GI toxicity ≥G2 were 13% (95% CI: 7-18%) and 2% (95% CI: 0-4%). For late GU and GI toxicity ≥G2 the pooled rates were 25% (95% CI: 14-35%) and 5% (95% CI: 1-9%). The pooled 2-year biochemical recurrence-free survival was 72% (95% CI: 64-92%).

**Conclusions:** SBRT in the re-irradiation of radiorecurrent prostate cancer is overall safe and effective also when applied with a C-arm Linac or an MR Linac. Further prospective data are warranted.

## 1. Introduction

Radiotherapy is one of the main treatment modalities of prostate cancer treatment for localized disease (1). However, up to a third of patients develop a recurrence after primary radiotherapy (2, 3). With modern imaging like PSMA-PET/CT, distinguishing between a local and regional recurrence or even metastatic disease is possible with high sensitivity and specificity (4-6). For patients with an isolated local recurrence, there are different treatment options like salvage surgery but also non-surgical options like high-intensity focused ultrasound (HIFU), cryotherapy and re-irradiation, with both brachytherapy and external beam radiotherapy (EBRT). A meta-analysis by Valle et al. found no significant difference for 2- and 5-year recurrence-free survival between radical prostatectomy and SBRT or brachytherapy in radiorecurrent prostate cancer. They did however find significantly lower rates of severe GU toxicity in patients treated with any form of radiotherapy (7).

Regarding EBRT, stereotactic radiotherapy (SBRT) has proven to be feasible with a promising oncological outcome and acceptable toxicity rates not only after EBRT but also brachytherapy (8).

Early series on prostate re-irradiation using SBRT date back more than 10 years (9, 10). In 2019, a large Meta-analysis on non-surgical local therapies for recurrent prostate cancer by Ingrosso et al. showed good outcomes for biochemical and local control as well as incontinence rates when using EBRT as salvage therapy. In this analysis only six out of the eight EBRT studies used SBRT for all patients in the re-irradiation setting (8). There was another systematic review in 2021 by Munoz et al. including studies on prostate re-irradiation using mostly brachytherapy but also EBRT or SBRT until 2019. They found acceptable biochemical failure rates with pooled 2- and 4-year BF rates of 24 % and 35.6 % and pooled high grade (≥ G3) late toxicity was 8.7 % (11).

Noticeably, earlier studies on SBRT for radiorecurrent prostate cancer were often done using a Cyberknife treatment machine (9, 10, 12-16). In the last years, however, there has been increasing evidence on prostate re-irradiation using a C-arm LINAC or a MR LINAC (17-33).

We therefore conducted this Systematic Review and Meta-Analysis on prostate re-irradiation using SBRT including studies published from 2020 to 2023 to serve as an update of the above-mentioned Meta-analyses.

## 2. Materials and Methods

### 2.1 Study search and selection process

The PICO criteria (Population, Intervention, Control, Outcome) were used for this Systematic Review with the following criteria: population – patients with radiorecurrent prostate cancer after primary radiotherapy; intervention – SBRT to radiorecurrent cancer in the prostate; control - historical controls from published phase II/III studies; outcome - rate of acute and late toxicities after SBRT and b) biochemical control after SBRT (34, 35).

This analysis was done in accordance with the Preferred Reporting Items for Systematic Reviews and Meta-Analyses (PRISMA) recommendations (36). We searched the Pubmed and Embase databases with the following full-text queries in October 2023: “repeat” AND “radiotherapy” AND “prostate”, “re-irradiation” AND “prostate”, “re irradiation” AND “prostate”, “reirradiation” AND “prostate”, “re treatment” AND “prostate” AND “SBRT”, “retreatment” AND “prostate” AND “SBRT”. As for publication date, we applied a filter for the years 2020-2023. There was no language limitation. All initially identified records were copied to an Excel sheet (Microsoft Cooperation, Redmond, WA, USA), which was used to automatically identify and remove duplicates. Further manual removal of duplicates was done. Out of the initially identified records, only full-text articles in English reporting primary data were included in the further process. For review articles, opinions, etc. the references were checked to identify any further records that had not been identified, yet. For cross reference, also terms like “extreme hypofractionation” or “ultra-hypofractionation” were considered. As the next step, only papers reporting data on prostate re-irradiation using SBRT and re-irradiation to the prostate in at least a part of the cohort were selected. Mixed cohorts including patients receiving re-irradiation to the prostate bed were allowed. However, to identify the final papers included in this analysis, papers without independent reporting of the outcome in patients with SBRT after primary radiotherapy were excluded. The identification and selection process was done twice by two of the co-authors independently (CS and RF). A third co-author served as the final judge as to which papers were included (PW). Quality assessment of the included publications was done using a modified Delphi tool for case-series studies (8, 37). The results are shown in table S1 in the supplement.

**Table S1.**
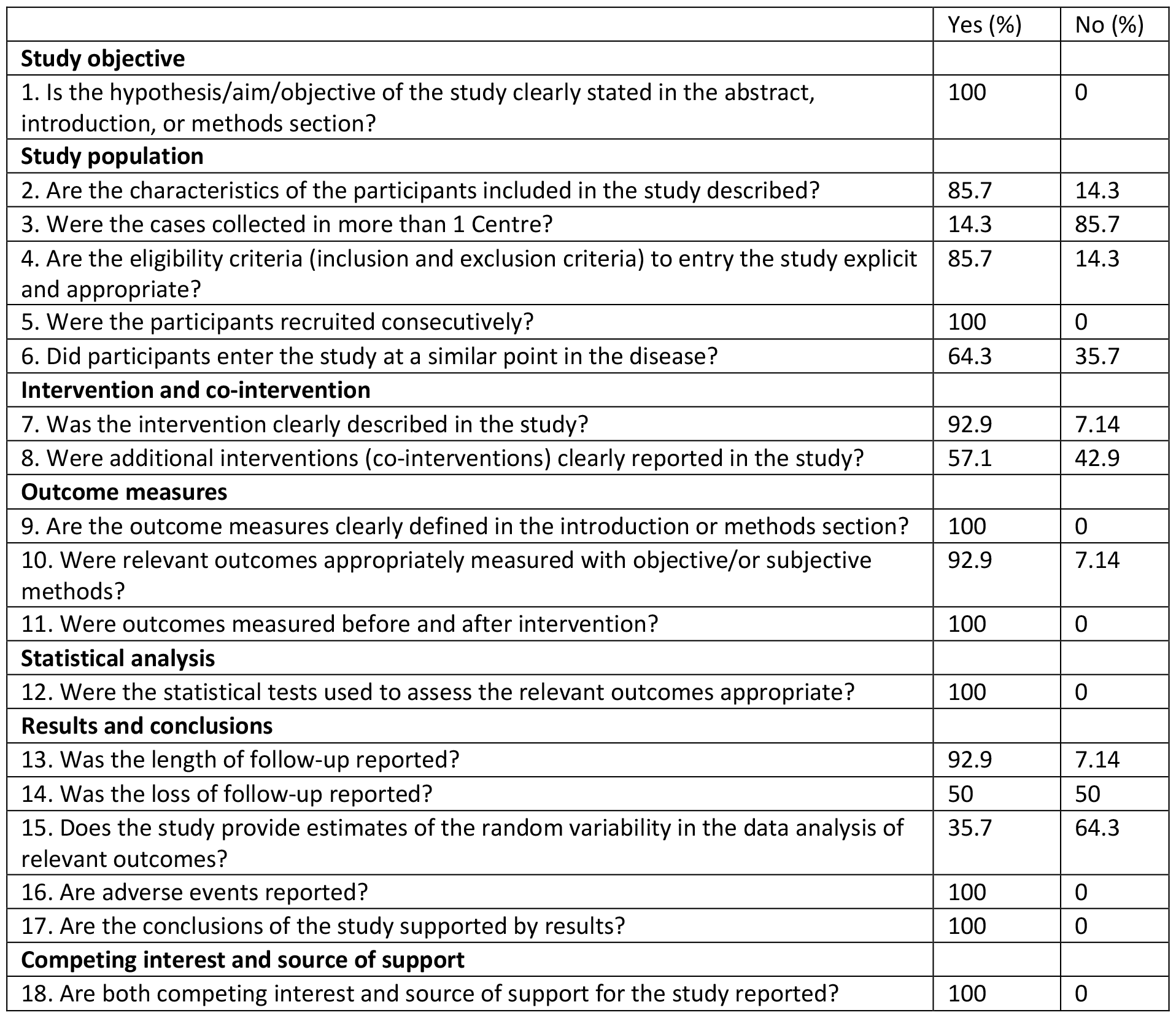
Results of the modified Delphi technique (n = 14, 100%)

|  | Yes (%) | No (%) |
| --- | --- | --- |
| **Study objective** |  |  |
| 1. Is the hypothesis/aim/objective of the study clearly stated in the abstract, introduction, or methods section? | 100 | 0 |
| **Study population** |  |  |
| 2. Are the characteristics of the participants included in the study described? | 85.7 | 14.3 |
| 3. Were the cases collected in more than 1 Centre? | 14.3 | 85.7 |
| 4. Are the eligibility criteria (inclusion and exclusion criteria) to entry the study explicit and appropriate? | 85.7 | 14.3 |
| 5. Were the participants recruited consecutively? | 100 | 0 |
| 6. Did participants enter the study at a similar point in the disease? | 64.3 | 35.7 |
| **Intervention and co-intervention** |  |  |
| 7. Was the intervention clearly described in the study? | 92.9 | 7.14 |
| 8. Were additional interventions (co-interventions) clearly reported in the study? | 57.1 | 42.9 |
| **Outcome measures** |  |  |
| 9. Are the outcome measures clearly defined in the introduction or methods section? | 100 | 0 |
| 10. Were relevant outcomes appropriately measured with objective/or subjective methods? | 92.9 | 7.14 |
| 11. Were outcomes measured before and after intervention? | 100 | 0 |
| **Statistical analysis** |  |  |
| 12. Were the statistical tests used to assess the relevant outcomes appropriate? | 100 | 0 |
| **Results and conclusions** |  |  |
| 13. Was the length of follow-up reported? | 92.9 | 7.14 |
| 14. Was the loss of follow-up reported? | 50 | 50 |
| 15. Does the study provide estimates of the random variability in the data analysis of relevant outcomes? | 35.7 | 64.3 |
| 16. Are adverse events reported? | 100 | 0 |
| 17. Are the conclusions of the study supported by results? | 100 | 0 |
| **Competing interest and source of support** |  |  |
| 18. Are both competing interest and source of support for the study reported? | 100 | 0 |

### 2.2 Data Extraction Process

The following data were extracted from the included manuscripts: first author, year of publication, journal, study design (retrospective, prospective), study period, overall number of patients included, number of patients included after definite radiotherapy, radiation treatment technique at first irradiation, total treatment dose at first irradiation, treatment machine, diagnosis criteria for recurrence, number of patients with biopsy at recurrence, defined minimum interval between RT courses, median time between RT courses, median follow up, total radiation treatment dose, Isodose line (IDL), target of treatment (entire prostate vs. focal vs. mixed), scheduling (alternating vs. consecutive days vs other), staging at re-irradiation, number of patients receiving ADT at re-irradiation, rates of acute and late toxicities (according to the Radiation Therapy Oncology Group (RTOG) or Common Terminology Criteria for Adverse Events (CTCAE) classification), data on biochemical control, data on target delineation for SBRT. The data were extracted by two independent co-authors (CS and RF) with third co-author serving as the referee in case of potential disagreements (PW).

### 2.3 Statistical Analysis

All statistical analyses were performed using R (v. 4.3.1) and RStudio (v. 2023.09.1+494) with the robometa (v. 2.1), metafor (v. 4.4.0) and dpyler (v. 1.1.3) libraries.

Following heterogeneity testing using I2, a random effects model with a restricted maximum likelihood estimator (38) was used for estimating the combined effect. Funnel plot asymmetry was assessed visually and using Egger’s test to estimate the presence of publication and/or small study bias (39).

## 3. Result

### 3.1 Selected studies

We identified a total of 1294 studies from the initial search of the databases. From these records, 677 duplicates were removed. From the resulting 617 records, 295 records were removed due to no available full text, no record in English language or no recording of primary data being present. Of the remaining 322 records used for screening, another 284 were excluded because no data on re-irradiation after initial treatment for PCA and/or no outcome data was reported, resulting in 38 records. In the final step, 14 papers included data on patients treated with SBRT for radiorecurrent prostate cancer after primary radiotherapy and were selected for the Systematic Review while 24 papers were removed during this step (23-33, 40-42). Figure 1 shows the consort diagram of the study selection process.

**Figure 1.**
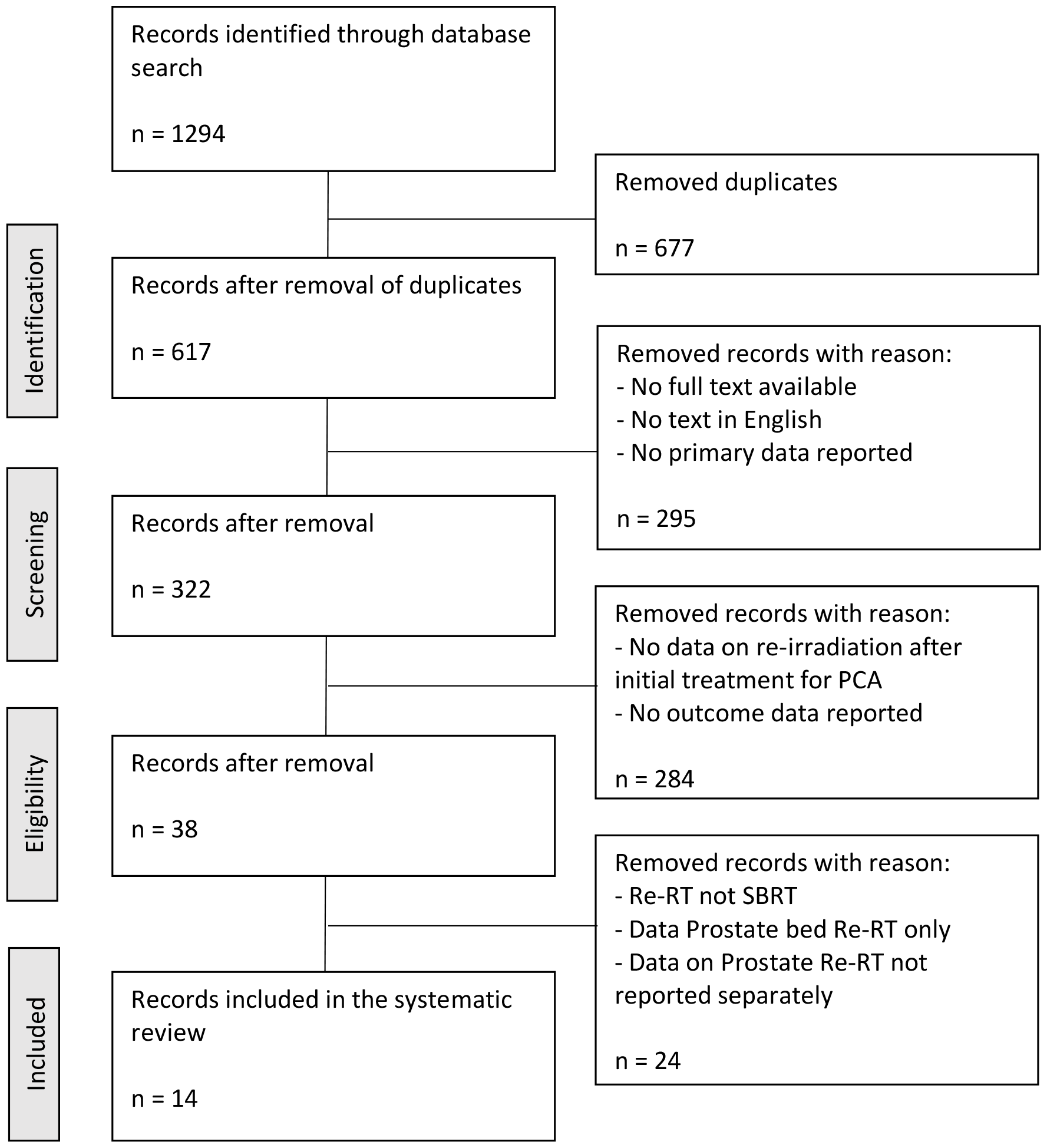
Paper selection process

Among the 14 selected papers, there were five prospective analyses, seven were retrospective analyses, one retrospective analysis of a prospective database and one was a small case series. Two publications included patients with re-irradiation to the prostate or the prostate bed and separate reporting for both groups. Table 1 shows an overview of the included publications.

### 3.2 Target volume and prescription dose

There were differences regarding both, target volume delineation and dose prescription between the included publications. In five publications, focal re-irradiation was done while whole prostate re-irradiation was used in four publications. Another five publications included cohorts with mixed focal or whole prostate re-irradiation. The most common dose concepts were 5 × 6 Gy, 5 × 7 Gy or 6 × 6 Gy given either on alternating or consecutive days. Further details on target delineation and dose prescription can be found in table 2.

**Table 2.** Target delineation and dose concepts.

| Author/year | Dose | IDL | Scheduling | Target |
| --- | --- | --- | --- | --- |
| Allali et al./2023 (40) | 5 x 6 Gy (22%) or 5 x 7 Gy (5%) or 6 x 6 Gy (73%) | 90% | alternating days | CTV = Prostate (incl seminal vesicles where necessary)<br>PTV = CTV + 3-5 mm |
| Matrone et al./2021 (29) | 5 x 7 Gy | homogenous | consecutive days | GTV = lesion<br>CTV = GTV + 3mm<br>PTV = CTV + 3mm |
| Greco et al./2022 (27) | 5 x 7 Gy (60%) or 5 x 8 Gy (13.4%) or 7/8 Gy SIB (26.6%) | homogenous | consecutive days (87%) | CTV = CTV = Prostate (93%) or partial prostate (7%)<br>PTV = CTV + 2 mm |
| Bergamin et al./2020 (24) | 6 x 6 Gy (72%) or 6 x 6.33 Gy (28%) | homogenous | alternating days | GTV = lesion<br>CTV = GTV + 3 mm<br>PTV = CTV + 3 mm |
| Lewin et al./2021 (28) | median 5 x 6.5 Gy (range, 6 - 8 Gy) | not reported | alternating days | CTV = lesion (43%) or prostate (57%)<br>PTV = CTV + 3-5 mm |
| Pasquier et al./2023 (33) | 5 x 6 Gy (38%) or 6 x 6 Gy (57%) or 5 x 5 Gy (5%) | not reported | alternating days | GTV = lesion<br>CTV = GTV + 5-7 mm<br>PTV = CTV + 2mm |
| Cozzi et al./2023 (25) | 5 x 6 Gy | homogen | alternating days | CTV = lesion (90%) or prostate (10%)<br>PTV = GTV + 5-6 mm |
| Ozyigit et al./2020 (32) | 5 x 6 Gy | not reported | alternating days | GTV = lesion<br>PTV = GTV + 1 mm |
| Cuccia et al./2022 (26) | 5 x 6 Gy | homogenous | alternating days | CTV = prostate<br>PTV = CTV + 3-5 mm |
| Montalvo et al./2022 (30) | 5 x 7 Gy (20%) or 5 x 8 Gy (40%) or 6/7Gy SIB (20%) | homogen | weekly | GTV = lesions<br>PTV not reported |
| Augugliaro et al./2021 (23) | median 5 (range, 2-10) x 6 Gy (range, 3 - 12 Gy) | not reported | not reported | GTV = lesion, CTV = prostate or lesion with margins<br>PTV = CTV + 5 mm (3 mm posterior), margin reduction to 1 mm in selected cases |
| Miszczyk et al./2023 (42) | 5 x 7.25 Gy (64%) or 5 x 6.75 Gy (7%) or 5 x 5.5 Gy (7%) or other (22%) | not reported | not reported | CTV = lesion (27%) or prostate (73%) PTV = CTV + 5 mm (3 mm posterior) |
| Fuller et al./2020 (41) | 5 x 6.8 Gy | "HDR like" | consecutive days | CTV = prostate plus any extension, no PTV |
| Nikitas et al./2023 (31) | 5 x 6- 6.8 Gy (some SIB with 7.5 Gy) | homogenous | not reported | CTV = prostate (incl seminal vesicles where necessary), PTV = CTV + 3-5mm (LINAC) or 3mm (MR LINAC); 4 mm to the seminal vesicles where applicable |

### 3.3 Acute and Late Toxicities

Toxicity scoring was done using the RTOG/EORTC toxicity scoring system in two publications (23, 29) and the CTCAE scoring system (versions 3 to 5) in nine publications (25-28, 31, 33, 40-42). In two publications, CTCAE was used for acute toxicity while RTOG was used for late toxicity (24, 32). The scoring system was not specified in one publication (30). Both toxicity systems overall have a moderate interscale agreement with more G1-2 toxicities being identified with the CTCAE scoring system (43, 44).

The rates of acute ≥ G2 GU and GI toxicities reported in the included studies range from 0.0 to 30.0 % and 0.0 to 25.0 % respectively. For late ≥ G2 GU and GI toxicity, those values are 4.0 – 51.8 % and 0.0 – 25.0 %. The rates for late ≥ G3 toxicity range from 0.0 % - 10.7 % for GI and 0.0 % - 23.2 % for GU.

### 3.4 Meta-analysis of acute toxicity

While the calculated I^2^ statistics for acute GU and GI toxicity greater than or equal to grade 2 did not indicate a high degree of heterogeneity (0 and 39.9%, respectively), the confidence intervals were wide due to the limited number and size of the included studies (95% CI: 0% – 85.4% and 0% - 79.7% respectively).

The pooled rate of acute GU toxicity greater than or equal to grade 2 was 13% (95% CI: 7% - 18%) and the pooled rate of acute GI toxicity greater than or equal to grade 2 was 2% (95% CI: 0% - 4%).

Egger’s test found significant funnel plot asymmetry for both acute GU toxicity greater than or equal to grade 2 (p = 0.007) and acute GI toxicity greater than or equal to grade 2 (p = 0.02) with larger studies reporting lower rates of toxicity.

The Forest plots are shown in figure 2 a) and b) and the associated Funnel plots as figures S1 a) - b) in the supplement.

**Figure S1.**
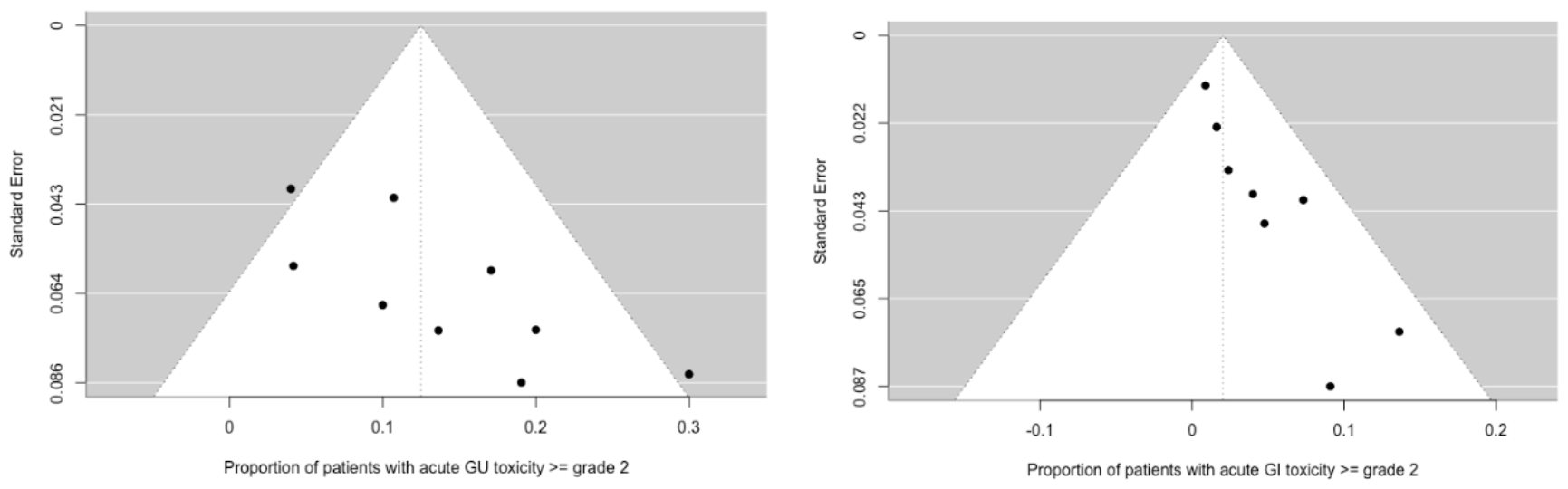
a) and b) Funnel Plots of the included publications for acute >=G2 GU (a) and GI (b) toxicity

**Figure 2.**
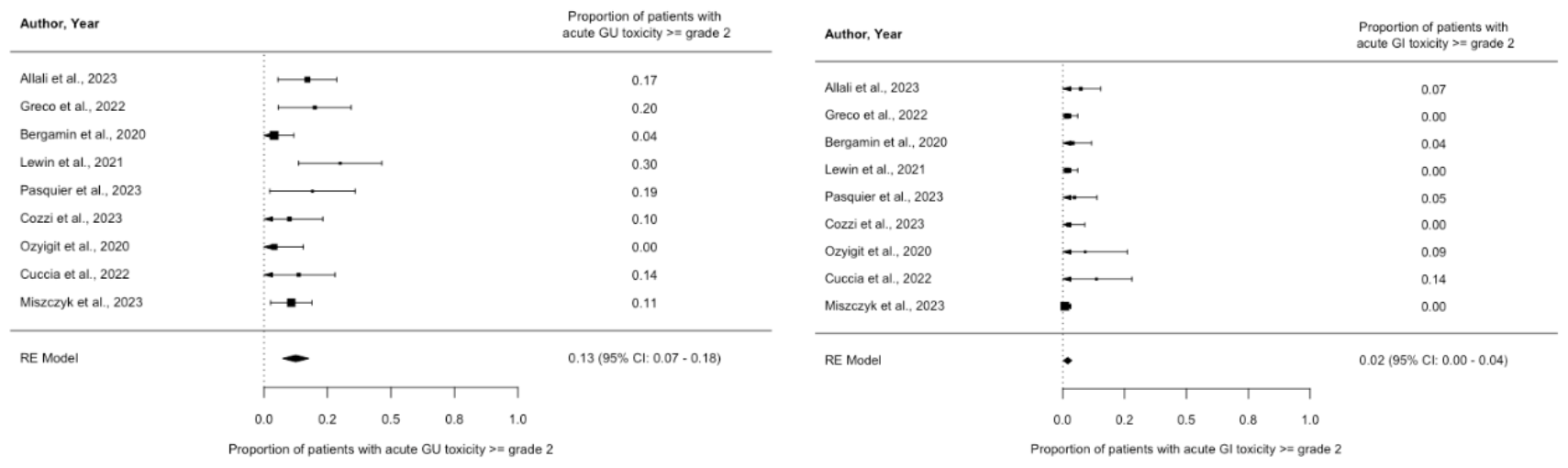
a) and b) Forest Plots of the included publications for acute >=G2 GU (a) and GI (b) toxicity

### 3.5 Meta-analysis of late toxicity

The calculated I^2^ statistics for late GU and GI toxicity greater than or equal to grade 2 were 77.9% (95% CI: 49.5% - 94.1%) and 68.4% (95% CI: 19.2% - 94.9%), respectively.

The pooled rate of late GU toxicity greater than or equal to grade 2 was 25% (95% CI: 14% - 35%), and the pooled rate of acute GI toxicity greater than or equal to grade 2 was 5% (95% CI: 1% - 9%).

Egger’s test did not find significant funnel plot asymmetry for late GU toxicity greater than or equal to grade 2 (p = 0.097). However, there was significant funnel plot asymmetry for late GU toxicity greater than or equal to grade 2 (p = 0.0007) with larger studies reporting lower rates of toxicity. The Forest plots for late toxicity are shown as figures 3 a) and b). The repective Funnel plots can be found as figures S2 a) and b) in the supplement.

**Figure S2.**
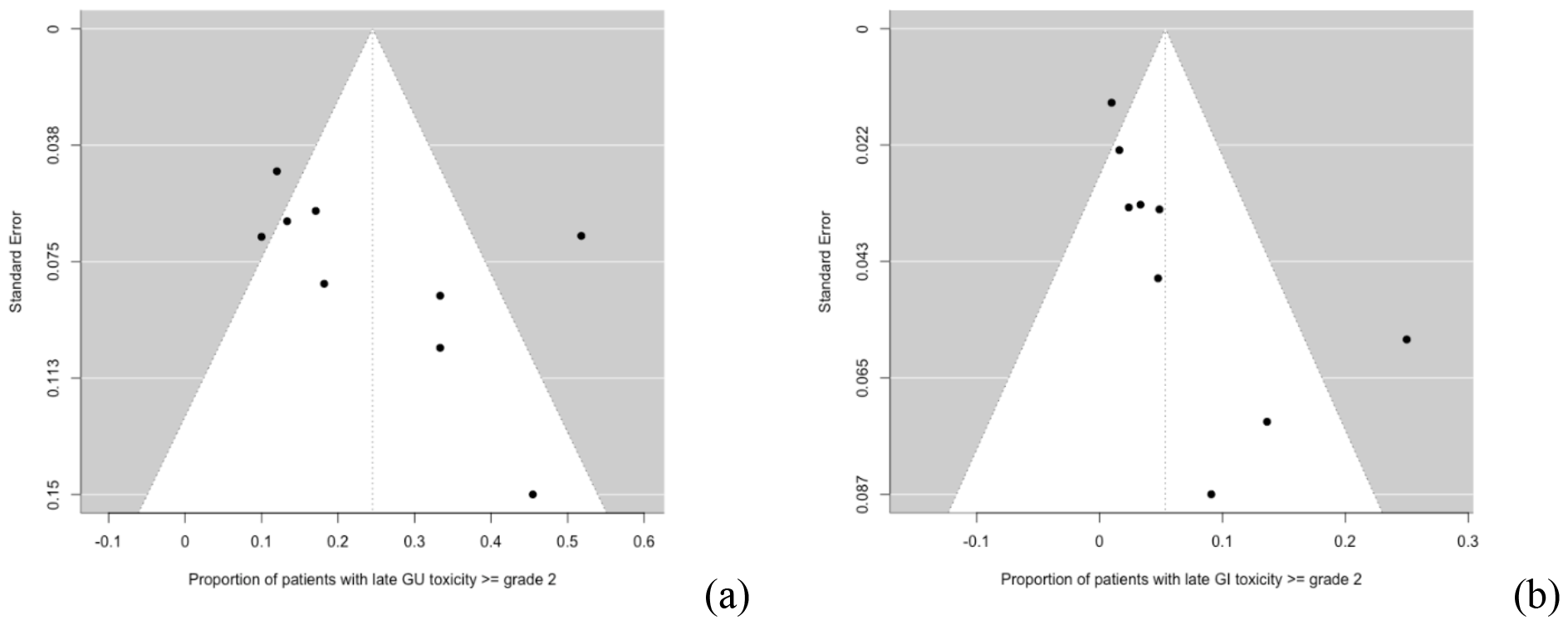
a) and b) Funnel Plots of the included publications for late >=G2 GU (a) and GI (b) toxicity

**Figure 3.**
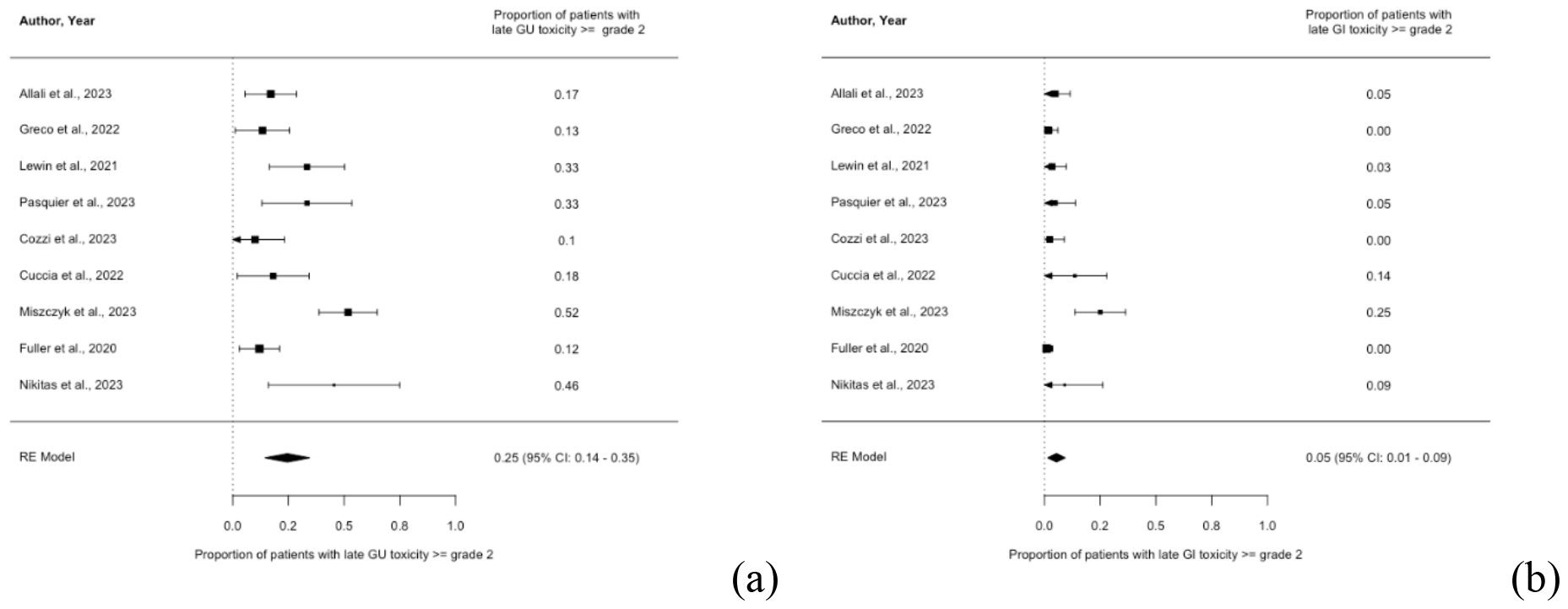
a) and b) Forest Plots of the included publications for late >=G2 GU (a) and GI (b) toxicity

Only few studies reported on factors associated with the occurrence of toxicity. Miszsczyk et al. found an association with G3+ toxicity for PTV size, the extent of salvage radiotherapy (focal vs. whole prostate) and the use of ADT in the univariate analysis of which PTV size and ADT remained significant in the multivariate analysis (42). For G3+ GU toxicity, Fuller et al found a significant difference for patients receiving EBRT as first radiotherapy versus more intensive first therapies (41). Greco et al. reported an association of G2 GU toxicity and gland volume (27) in univariate analysis.

### 3.6 Biochemical control

Data on 2-year BRFS was reported in 10 studies (23-25, 27, 29, 31, 32, 40-42). 2-year BRFS in those studies ranged from 48 % to 91 %. Median follow-up in those studies ranged from 19 months to 47.7 months.

### 3.7 Meta-analysis of biochemical control

The calculated I^2^ statistic for 2-year BRFS was 65.5% (95% CI: 26.3% - 91.8%). The pooled 2-year BRFS was 72% (95% CI: 64% - 92%). Egger’s test did not find significant funnel plot asymmetry (p = 0.135). Figures 4 shows the Forest plot for the 2-year bRFS and the Funnel Plot is shown as figure S3 in the supplement.

**Figure S3.**
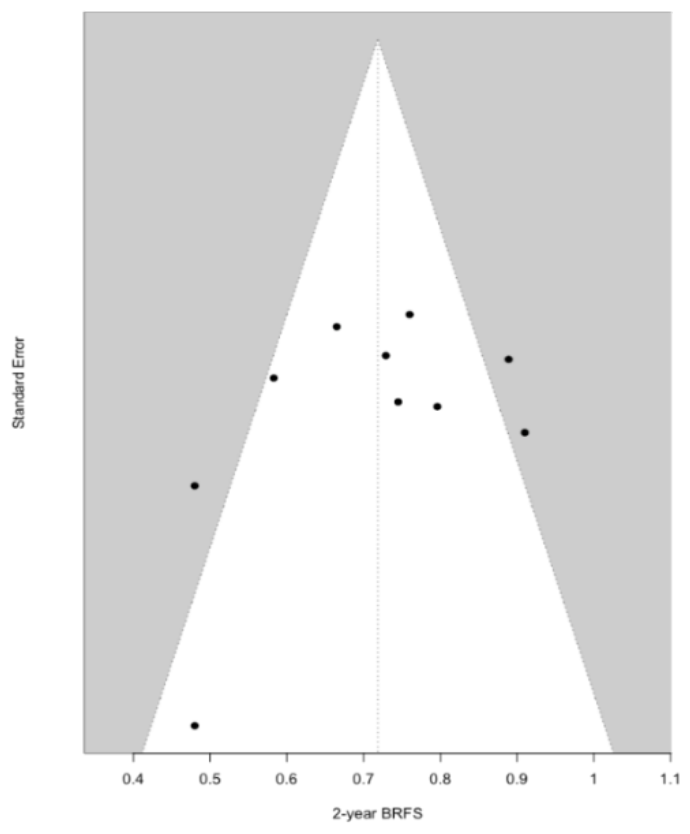
Funnel plot for 2-year bRFS

**Figure 4.**
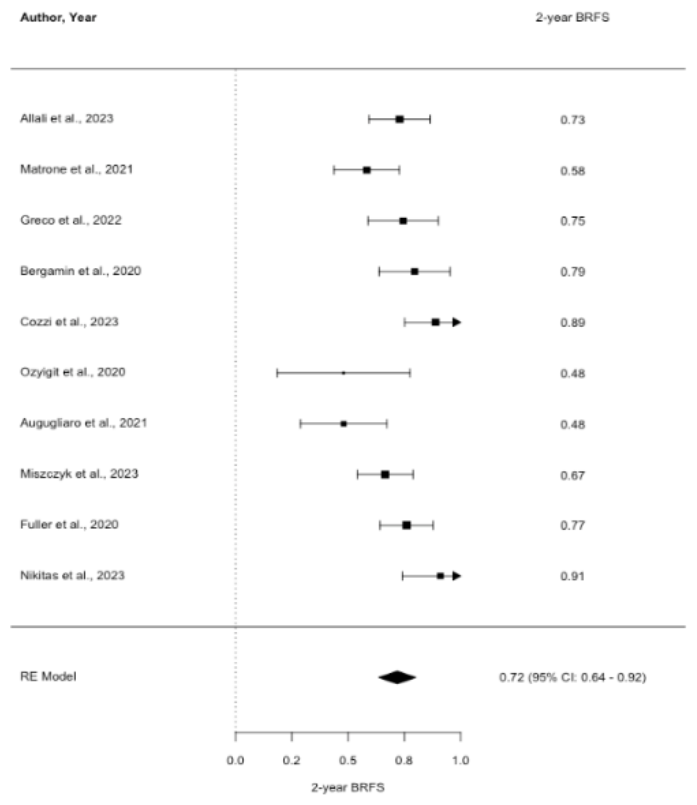
Forest plot for 2-year bRFS

Several factors associated with the biochemical outcome were reported in the included studies. Matrone et al. reported favourable outcomes in patients with a longer interval between the RT courses, a higher BED at first RT and the use of ADT at re-irradiation. Greco et al. identified the treatment modality at first RT (EBRT vs. brachytherapy) as a significant factor (27). Other factors that were associated with biochemical outcome were ISUP risk group at first RT and pre-salvage PSA level (41, 42).

However, Ozyigit et al. found no association for the use of ADT, primary RT dose, pre-salvage PSA or treatment machine and Greco et al. found no association for the interval between the RT courses, NCCN risk group at first RT or the use of ADT at re-irradiation (27, 32).

## 4. Discussion

After primary radiotherapy up to a third of patients develop a recurrence after primary definite radiotherapy (2, 3).

Generally, PSA bounces after definite radiotherapy are common, especially in the first years after radiotherapy (45-47). Biochemically failure was defined at the RTOG-ASTRO Phoenix Consensus Conference as a PSA rise above 2 ng/ml above nadir (48). In case of biochemical recurrence, PET CT scans and MRI are useful in distinguishing between a localised recurrence and nodal or metastatic recurrence as well as localization of the intraprostatic recurrence (4, 49-51). Biopsies to confirm and characterize the recurrence should be highly considered (52, 53). At the time of recurrence, about 10 % of patients present with localized intraprostatic recurrence (3, 54). In those cases, there are surgical and non-surgical approaches as potentially curative treatment (7). A Meta-analysis by Valle et al. found no difference in 2-year or 5-year recurrence-free survival between radical salvage prostatectomy compared to SBRT, LDR brachytherapy or HDR brachytherapy. They did however find significantly lower rates of severe (defined as ≥ G3) GU toxicity for any type of radiotherapy and for HDR brachytherapy also significantly lower rates of severe GI toxicity (7).

Generally, non-surgical approaches for radiorecurrent prostate cancer include high-intensity focused ultrasound (HIFU), cryotherapy, normofractionated external beam radiotherapy, SBRT, and HDR or LDR brachytherapy (8). Both brachytherapy and external beam radiotherapy were found to be good treatment options for locally recurrent disease. The latter is nowadays mostly done using SBRT, which next to technical advantages might also have biological advantages due to the low alpha/beta value of prostate cancer of approximately 1.5 (55, 56).

SBRT as salvage therapy for radiorecurrent prostate cancer has been used for more than 15 years (9, 10, 12). Early case series were usually done with a Cyberknife treatment machine whereas newer studies included cohorts treated with C-arm Linacs or MR Linacs (10, 12-17, 19, 22-33, 40-42, 57-63). Of the 14 studies included in this meta-analysis, only three included patients that were exclusively treated on a Cyberknife (40-42).

There is no consensus on target delineation, fractionation or scheduling (alternating vs. consecutive days). Regarding treatment dose, common fractionation schemes used in the included studies are 5 × 6 – 7.25 Gy or 6 × 6 Gy. With an estimated alpha/beta value of 1.5 for prostate cancer this results in EQD2 doses of 64 – 90 Gy_1.5_ and 38 – 49 Gy_3_ with an estimated alpha/beta value of three for organs at risk. In nine of the included studies, multiple dose fractionation schemes were used which further limits the possibility of making statements on the dose-response relationship. Target delineation in the included studies was also quite different ranging from focal to whole prostate re-irradiation, even including the seminal vesicles where necessary. Also for target delineation, three studies included mixed cohort that received either focal or whole gland irradiation.

Regarding toxicity, there is overall a large range of the reported acute and late GU and GI toxicity. The reported acute ≥G2 GU and GI toxicities range from 0 – 30 % and 0 – 25 %. This is well within the range of older data on acute toxicity after re-irradiation (9, 10, 13-16, 19, 62, 64, 65). In the meta-analysis, it is evident that despite the outliers with high toxicity range, the overall reported ≥G2 acute toxicity is rather low with 13 % for GU (95% CI 7 – 18%) for GU and 2% (95% CI 0 – 4%) for GI. Also, for late toxicity the overall range of ≥G2 toxicity is rather large with 4 – 51.8 % for GU and 0 – 25 % for GI.

It has to be noted that only one included publication reported data on a brachytherapy-only cohort at first RT (31). Eight studies mixed cohorts but usually low amount of brachytherapy patients (< 20%) except for Greco et al. where 43 % of patients had initial brachytherapy (37% LDR and 7% EBRT with LDR boost) (24, 26-28, 30, 31, 40-42). There is another study on mildly hypofractionated external re-irradiation after LDR showing low toxicity rates with 30% late G1 toxicity (66). More intensive first treatment was a risk factor for higher G3+ toxicity reported by Fuller et al. However, next to the 5 patients with initial brachytherapy this also included a patient with RP and a patient with SBRT (41). Contrary to this, Pasquier et al. found no association of toxicity and kind of first treatment (EBRT vs. brachytherapy) (67).

There however seems to be higher reported toxicity rates for whole gland re-irradiation versus focal irradiation, especially for GI (0 - 4.7% for focal vs. 0 – 25% for whole gland). PTV size was one of the factors associated with the risk of G3+ toxicity reported by Miszczyk et al. (42). However, a large retrospective analysis of the GETUG including 100 patients found no differences in toxicity depending on the treated volume (whole-prostate vs partial SBRT) or PTV (67).

As for the efficacy of SBRT in the re-irradiation setting of radiorecurrent prostate cancer, the 2-year bRFS rates reported in the ten studies included in the meta-analysis range from 48% to 91% (23-25, 27, 29, 31, 32, 40-42). In the random effects model the 2-year bRFS was 72 % (95% CI 64 – 92%). This seems to be overall consistent with older data (10, 13-15, 64, 67).

It has to be noted though that two studies included in the meta-analysis included node-positive or oligometastatic patients at re-irradiation. Greco et al included 13% node-positive and 10% oligometastatic patients and Miszczyk et al. even 19.6% oligometastatic patients. However, the reported 2-year bRFS of 74.5% and 66.5% were not the lowest of all included studies (27, 42). Also, Lewin et al. included node-positive (23.3%) and oligometastatic (3.3%) patients but reported no 2-year bRFS (28).

Although this systematic review and meta-analysis includes five prospective studies, more prospective data is important. There are currently the results of several trials awaited, among others the phase II of the GETUG AFU 31 for which results of phase I were included in this meta-analysis (33, 68). Additionally, a randomized comparison of salvage options in the radiorecurrent setting would be interesting, for example between brachytherapy and SBRT.

## 5. Conclusion

SBRT in the re-irradiation of radiorecurrent prostate cancer is overall safe and effective also when applied with a C-arm Linac or an MR Linac. Especially regarding the potential benefit of using an MR Linac, long-term outcome data is warranted.

## Data Availability

All data produced in the present study are available upon reasonable request to the authors.

